# Free-text MAUDE narratives provide a source-robust representation layer for biomaterial-device surveillance

**DOI:** 10.64898/2026.05.03.26352339

**Authors:** Hongyi Chen

## Abstract

Implantable biomaterial devices require effective post-market surveillance because clinically important failure patterns often emerge only after widespread use. However, surveillance workflows often rely on structured coded summaries that compress heterogeneous adverse-event narratives into coarse categories. This study compares coded and free-text narrative representations across 1,500 FDA MAUDE reports from three biomaterial device classes (coronary stents, bone cement, and surgical mesh) to test whether narratives preserve a more source-robust surveillance representation. Under manufacturer-held-out evaluation, narrative TF-IDF features outperformed structured code-only features (macro F1 0.925 versus 0.827), while delexicalized narratives retained strong grouped performance after masking device-class, manufacturer, brand, and legal-template tokens (F1 0.897). Narrative topics resolved reported events into procedural, anatomical, host-response, and reporting-context patterns, and an interpretable classifier recovered code-derived complication phenotypes from narrative text alone (mean F1 0.902, AUC 0.967). These findings support free-text adverse-event narratives as a complementary representation layer for post-market device surveillance, while remaining bounded by passive adverse-event reporting limitations and requiring validation across additional years, device classes, and independently adjudicated outcomes.

**Author Summary:** When an implanted medical device fails inside a patient, the event is reported to the FDA’s MAUDE database. Each report includes both a standardized code and a written narrative describing what happened. We asked whether these two representations carry the same information. Using 1,500 reports covering coronary stents, bone cement, and surgical mesh, we found that coded fields lose much of the clinical detail present in narratives. Importantly, narrative-based classifiers remained accurate even when tested on reports from manufacturers not seen during training, while code-based classifiers dropped substantially. This matters because real-world surveillance must generalize across different reporting sources. We also found that narrative text can recover clinically meaningful complication patterns that are defined by codes, and that most reports never name the specific biomaterial involved. These findings suggest that narrative text deserves a more central role in post-market device monitoring, complementing the coded fields that current surveillance pipelines rely on.

## Introduction

Implantable medical devices approved through the FDA’s 510(k) or premarket approval pathways may encounter failure modes that emerge only after widespread clinical deployment, making post-market surveillance essential [1,2]. High-profile device withdrawals and safety actions illustrate this pattern: metal-on-metal hip implants were linked to elevated revision rates detected years after approval [3,4], transvaginal mesh was reclassified and ultimately withdrawn from the U.S. market in 2019 [5], and drug-eluting coronary stents raised late-thrombosis concerns that reshaped antiplatelet therapy guidelines [6,7]. The challenge is particularly acute for biomaterial devices, where metals, polymers, ceramics, and composites can fail through diverse pathways involving corrosion, wear, fatigue, host-mediated inflammation, and tissue–device interaction [10–14]. A single bone-cement event may involve PMMA monomer toxicity, exothermic tissue damage, and fat embolization [8], while a mesh erosion event may reflect both material degradation and anatomical biomechanical factors [5]. Effective surveillance therefore depends on representing reported events in a way that preserves this clinically meaningful heterogeneity.

In the United States, the primary passive surveillance instrument is the FDA’s Manufacturer and User Facility Device Experience (MAUDE) database, which receives several hundred thousand adverse-event reports annually [1,9]. Each MAUDE report contains two parallel representations: a structured coded layer drawn from standardized problem vocabularies maintained by the International Medical Device Regulators Forum [10], and a free-text narrative written by the reporter. In practice, downstream surveillance workflows rely predominantly on the coded layer. However, this dependence creates a representation bottleneck: a detailed narrative describing anatomy, procedural context, device behavior, and clinical consequences can be reduced to one or two standardized labels. Evidence from drug safety supports the hypothesis that coded fields systematically underrepresent phenotypic complexity: Harpaz et al. showed that structured coding captures only a fraction of adverse-event signal present in clinical narratives [11], Luo et al. demonstrated that NLP consistently recovers pharmacovigilance signals missed by coded fields [12], and scalable deep-learning approaches have confirmed that unstructured text contains predictive information beyond what structured elements capture [13,14]. In the device domain, NLP applied to regulatory databases has similarly demonstrated richer phenotypic detail in free-text reports than in their structured counterparts [15,16]. These findings motivate direct comparison of coded and narrative MAUDE representations.

Prior MAUDE studies have applied machine learning and NLP to device-specific analyses, including cochlear-implant adverse-event classification [17], microwave-ablation report analysis with large language models [18], health-information-technology event classification [19], and narrative completeness assessment for analgesia devices [20]. Bala et al. demonstrated that classifiers applied to MAUDE narratives identify safety-relevant events with high accuracy [21], and Brown et al. revealed systematic miscategorization of fatal outcomes in MAUDE death reports [22]. Mishali et al. showed that reporting-source variation, specifically whether a report originates from a manufacturer, user facility, or voluntary reporter, meaningfully affects surveillance patterns and introduces systematic biases into coded fields [23]. However, three critical gaps remain unaddressed. First, no study has directly compared coded and narrative MAUDE representations within a multi-category biomaterial-device context [17,19,21,22]. Second, no study has tested whether narrative representations remain more transferable than coded taxonomies when the reporting source changes [21,23]. Third, the IDEAL-D framework and coordinated registry initiatives presuppose that data representations feeding surveillance systems preserve sufficient phenotypic resolution to detect meaningful safety signals [24,25], yet this assumption has not been empirically tested for MAUDE narratives versus codes.

This gap is addressed here using a biomaterial-device testbed spanning coronary stents, bone cement, and surgical mesh, three device categories in which material, procedural, anatomical, and host-response factors create heterogeneous reported failure patterns [26–30]. Three primary contributions are presented. First, report-level compression is quantified in the structured MAUDE problem-code layer. Second, it is tested whether narrative representations remain more portable than coded taxonomies under manufacturer-held-out reporting-source shift, including delexicalized sensitivity analysis. Third, it is assessed whether narrative text can recover clinically interpretable, code-derived complication phenotypes. As a secondary exploratory analysis, sparse material mentions in MAUDE narratives are compared with material visibility in PubMed and https://ClinicalTrials.gov to generate hypotheses for material-aware surveillance. The central hypothesis is that free-text narratives preserve a higher-resolution and more source-robust representation of reported biomaterial-device failures than coded fields alone.

## Results

### Study design and cohort

This section defines the cohort and the two surveillance representations evaluated throughout the study: structured coded fields and free-text narratives. A total of 1,500 adverse event reports were extracted from the MAUDE database via the openFDA API, comprising 500 reports each for coronary stents, bone cement, and surgical mesh from calendar year 2022. Event type proportions (67% Injury, 32% Malfunction, 1% Death; Fig 1A) reflect reporting patterns rather than true incidence rates. Narrative availability was 100%, and word count distributions (Fig 1B) revealed structural differences across device classes: stent narratives were longest (median 296 words), reflecting the procedural complexity of percutaneous coronary interventions; bone cement reports were intermediate (median 257 words), augmented by detailed manufacturer narratives; and mesh reports were shortest (median 158 words), consistent with more formulaic reporting templates. Summary statistics are shown in Fig 1C. Because the cohort was balanced by design at 500 reports per device class, all between-class comparisons describe representation behavior within a constructed analytic benchmark rather than differences in population-level device safety. These descriptive differences characterize the corpus, but the central question is whether the narrative layer preserves more portable failure information than the coded layer.

**Figure 1.**
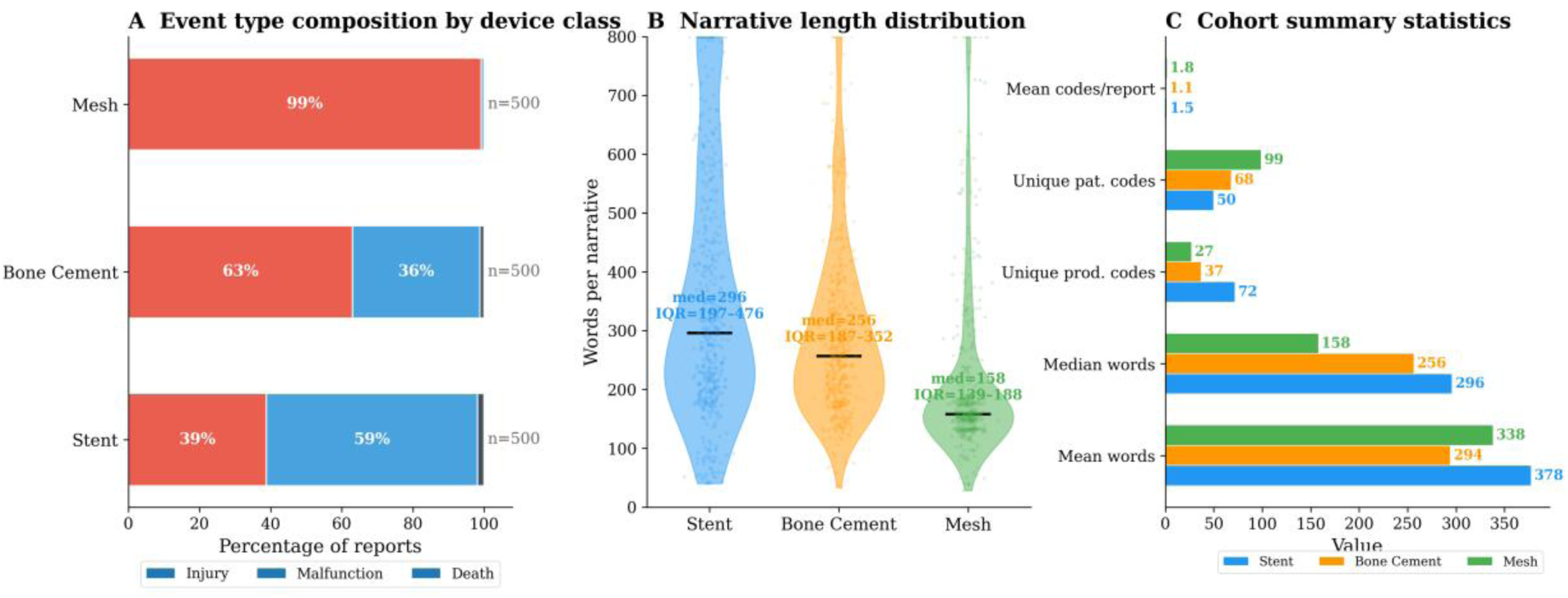
Benchmark cohort across three biomaterial device classes. (A) Event-type composition by device class; proportions describe reporting patterns rather than incidence. (B) Narrative word-count distributions by device class, with medians indicated. (C) Cohort-structure summary by device class, including mean and median narrative length, unique product and patient problem codes, and mean product codes per report. Together, these panels define the balanced benchmark and motivate comparison of coded and narrative surveillance representations.

### Structured codes provide a standardized but low-resolution surveillance baseline

The coded MAUDE layer is next characterized as a standardized baseline representation, testing how much report-level resolution it retains. Analysis of the coded taxonomy revealed device-specific frequency profiles (Fig 2A,B). Bone cement reports were dominated by “Loss of or Failure to Bond” and manufacturing-related codes; stent reports featured “Material Deformation” and “Failure to Advance”; mesh reports showed high rates of “Patient Device Interaction Problem” and “Defective Device.” Within-category normalization reveals distinct code profiles across device classes.

**Figure 2.**
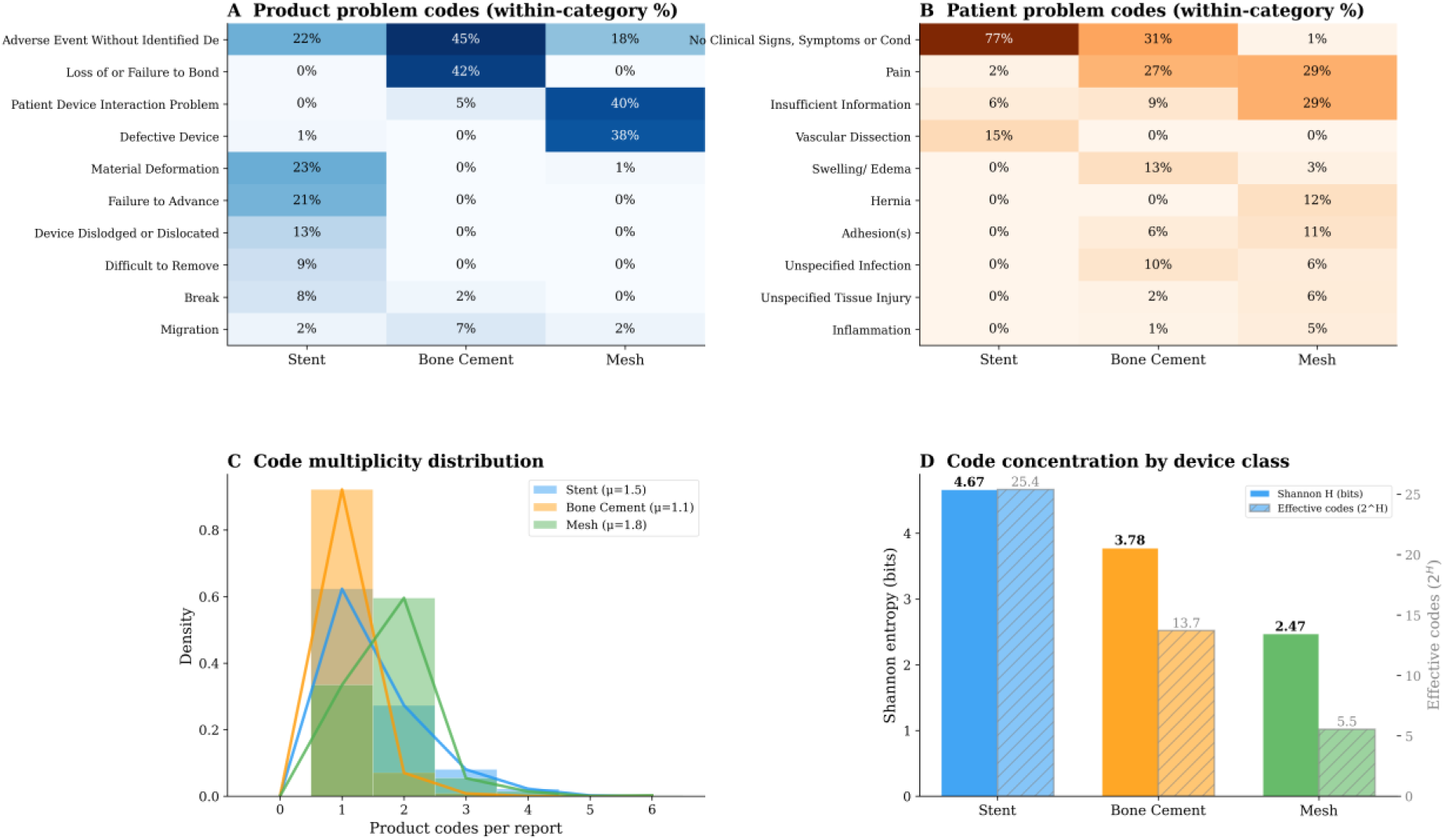
Coded surveillance provides a standardized but low-resolution baseline. (A) Within-category percentage heatmap of the dominant product problem codes, showing distinct device-side failure signatures across the three biomaterial device classes. Cell values and color intensity represent the share of reports within each device class that carry each code. (B) Within-category percentage heatmap of the dominant patient problem codes, revealing corresponding differences in reported clinical outcomes. (C) Overlapping density histograms of codes per report, showing that most events are summarized by only one or two product problem codes. (D) Shannon entropy and effective number of codes per device class, quantifying the concentration of code usage; low effective counts indicate that a narrow subset of codes dominates each category despite large nominal vocabularies.

However, most reports contained only 1–2 coded problems (Fig 2C), meaning that complex multi-mechanism events are compressed into minimal representations. A stent report describing a calcified-vessel deployment failure with guidewire entrapment and emergency surgical bailout receives the same single code as a routine malfunction. The effective number of commonly used codes (Fig 2D), estimated from Shannon entropy, was much smaller than each nominal vocabulary, indicating that a narrow practical code set dominates each device category despite large available vocabularies. Together, these results support the interpretation that the coded taxonomy provides a useful standardized baseline but that most reports are compressed into a small number of labels. An illustrative narrative-to-code compression example is provided in S4 to illustrate how multi-step procedural and clinical detail can collapse into a single product-problem label. This compression motivates the central question: does the narrative layer preserve the detail that coding discards? Extended code analyses are in S1–S3, S12, and S15.

### Narratives resolve coded events into higher-resolution clinical and reporting themes

The next question is whether free-text narratives resolve the same reports into richer clinical and reporting themes than those preserved by coded fields alone. TF-IDF vectorization (5,000 features, unigrams and bigrams, min_df = 3) extracted category-specific vocabularies substantially richer than any single code (Fig 3A). Stent narratives were dominated by cardiovascular procedural terms (catheter, lesion, balloon, deploy), bone cement by orthopedic fixation terms (cement, knee, hip, fixation), and mesh by hernia-related terms (hernia, recurrence, abdominal, erosion). These vocabularies encode procedural context, anatomical specificity, and temporal detail that the coded taxonomy cannot represent.

**Figure 3.**
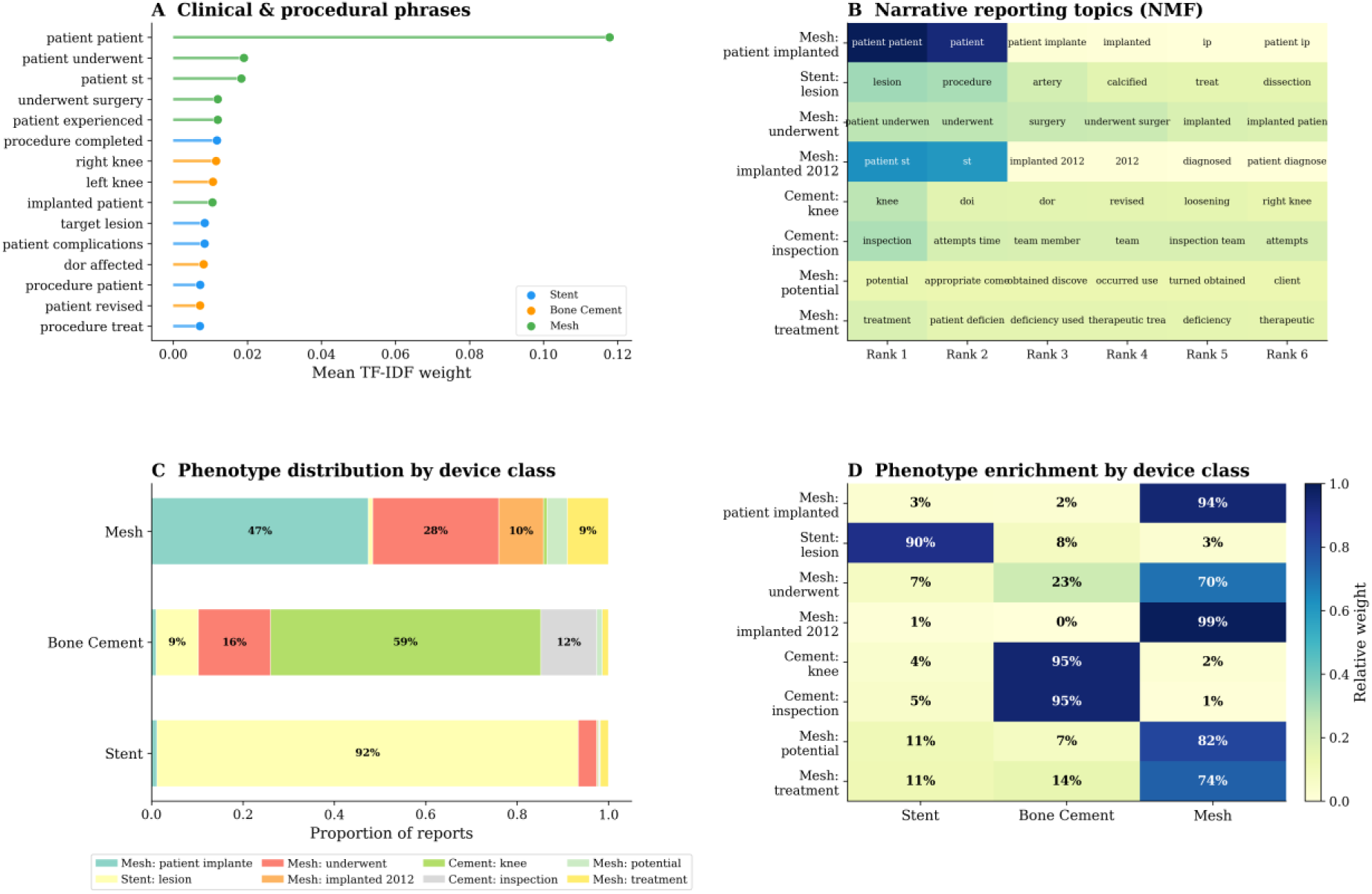
Narrative text resolves coded events into clinical and reporting themes. (A) Top multi-word features from cleaned event descriptions after removal of legal, manufacturer-template, and administrative boilerplate, colored by dominant device class. (B) NMF topic–term matrix showing eight latent narrative themes. (C) Theme distribution by device class. (D) Topic–device enrichment heatmap quantifying how each theme maps to specific device categories. Together, these panels show that narratives preserve device-specific procedural, anatomical, and complication-related information alongside reporting-pattern variation that is largely invisible in coded fields.

NMF topic modeling on the cleaned event description text (after removing legal, administrative, and manufacturer-template language; see Methods) resolved the narrative space into 8 reporting themes (Fig 3B). Several topics captured device-specific clinical content: a stent procedure topic dominated by coronary vocabulary (lesion, artery, calcified, dissection), a bone cement orthopedic topic (knee, revision, loosening, component), and a mesh surgery topic (underwent surgery, implanted, experienced pain). Other topics reflected reporting templates and legal language that persist even after preprocessing, concentrated in mesh reports. This mixture of clinical and reporting content is itself informative, indicating that the narrative signal is not uniformly clinical but includes device-specific regulatory patterns invisible in structured codes. Topic distribution by device class (Fig 3C) showed strong device–theme associations, and the enrichment heatmap (Fig 3D) quantifies this mapping. Topics were coherent and stable across random seeds (S4). For phenotype discovery, the event-description field was used after removing legal, administrative, and manufacturer-template language; for predictive representation benchmarks (Figs. 4–5), the full concatenated narrative representation described in Methods was used.

**Figure 4.**
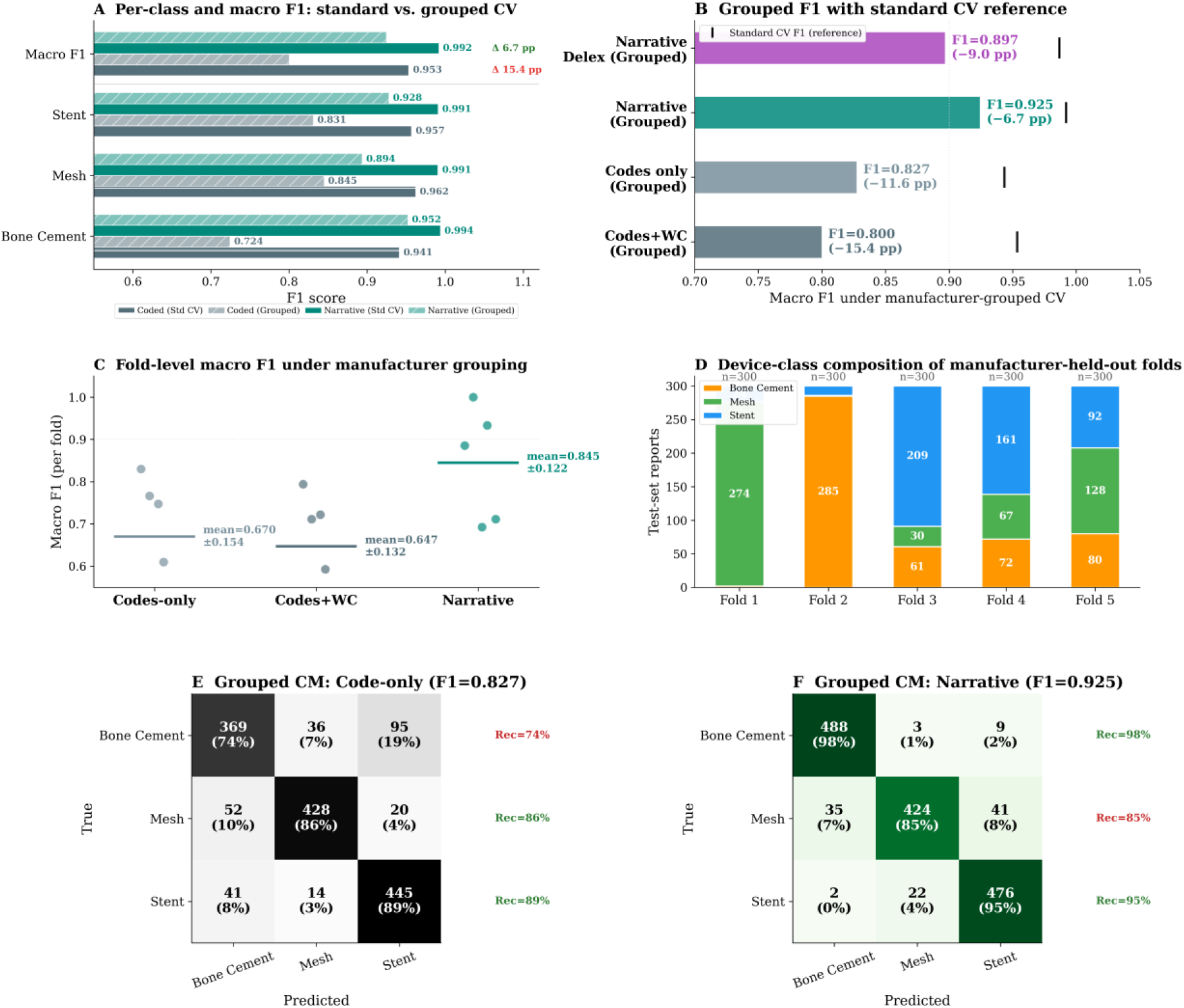
Narrative representations retain higher portability under manufacturer-held-out evaluation. Device-class prediction was used as a representation benchmark to compare coded and narrative features under reporting-source shift. (A) Standard and manufacturer-held-out macro F1 across the four representations. (B) Aggregate grouped macro F1 for structured codes plus word count, code-only structured features, original narrative TF-IDF, and delexicalized narrative TF-IDF. (C) Fold-level macro F1 values showing variability induced by manufacturer grouping. (D) Device-class composition of the five manufacturer-held-out folds. (E,F) Grouped confusion matrices for code-only and narrative representations. Because GroupKFold produces strongly imbalanced folds, aggregate grouped F1 should be interpreted together with fold-level variability and fold composition.

**Figure 5.**
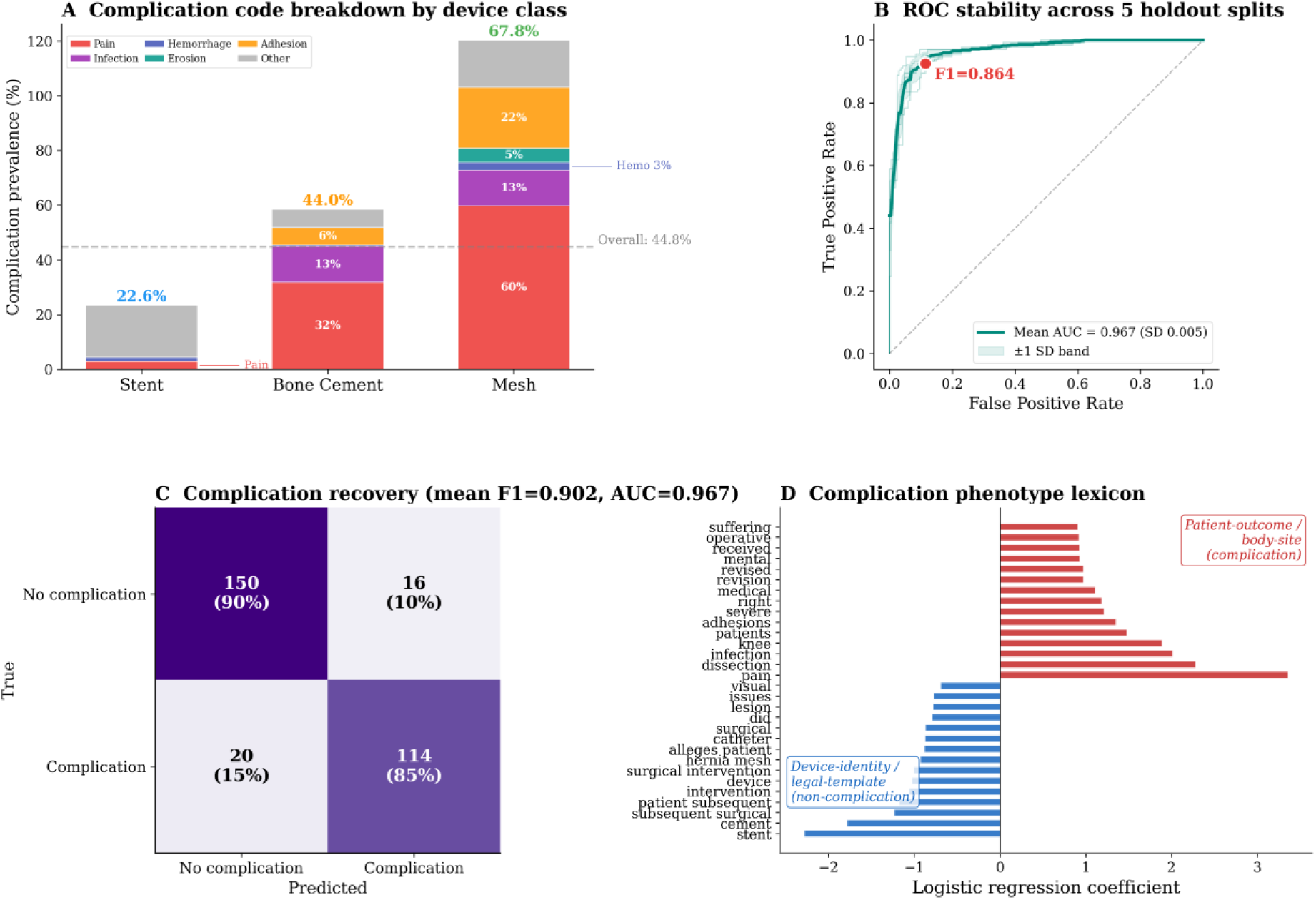
Narratives recover an interpretable code-derived complication phenotype. A binary complication phenotype was defined from clinically significant patient problem codes, yielding 672 positive reports among 1,500 total reports. (A) Complication prevalence by device class. (B) ROC curve for recovery of the code-derived complication phenotype from narrative TF-IDF features. (C) Confusion matrix from one representative holdout split; section-reported metrics are means across five repeated stratified holdouts. (D) Positive and negative logistic-regression coefficients, showing that complication recovery is driven by clinically interpretable patient-outcome and anatomical terms rather than device-identity or reporting-template language.

Narratives do not merely restate the coded taxonomy; they refine it into a phenotype space spanning procedure, anatomy, host response, and reporting context. While the topic layer does not yield uniformly clinical phenotypes, the analyses collectively show that narrative text carries richer and more heterogeneous information than the coded taxonomy, spanning device-specific clinical content and reporting-pattern variation invisible in structured codes. A t-SNE visualization of the TF-IDF narrative embeddings (S11) shows that device-class clusters are well separated in narrative feature space.

### Narrative representations remain more portable across manufacturers than coded taxonomies

The preceding analyses show that narratives carry richer information than codes; the central question is whether this advantage persists when the reporting source changes. Under manufacturer-grouped cross-validation, narrative features achieved higher aggregate manufacturer-held-out performance than the code-only representation (macro F1 0.925 versus 0.827). Adding narrative word count to the structured representation improved standard cross-validation slightly but reduced grouped performance (grouped F1 0.800), consistent with reporting-source verbosity acting as a portability confound. Because the study tests coded taxonomies against narrative text, the code-only model is the primary structured comparator, while the codes-plus-word-count model is retained as a sensitivity analysis. Manufacturer grouping induced substantial device-class imbalance across folds, so grouped point estimates should be interpreted as a source-shift stress test rather than as exact transportability effect sizes. Fold-level narrative macro F1 was variable (mean 0.845 ± 0.122), but narrative models outperformed structured models in 4 of 5 folds. The fold composition itself is informative: some held-out manufacturer folds were dominated by a single device class, reflecting manufacturer–device specialization in the reporting corpus.

To test whether the narrative advantage depended only on obvious identity tokens, the grouped benchmark was repeated after masking device-class keywords, manufacturer names, brand names, and legal-template tokens, which together represented 2.9% of all tokens (S3). Delexicalized narratives retained strong grouped performance (F1 0.897; S5), supporting the interpretation that the portability advantage is not driven solely by device, manufacturer, brand, or template-specific lexical cues.

For calibration, three-way device classification under standard 5-fold stratified cross-validation showed that both representations carry strong category signal: TF-IDF features achieved macro F1 = 0.992 (95% CI: 0.987–0.996) with logistic regression, compared to F1 = 0.943 (0.932–0.955) for gradient boosting on code-only features (S2). The gap between representations is small under standard conditions; the source-shift setting is what separates them. The principal advance is not higher classification accuracy alone, but the demonstration that narrative-derived representations remain more informative than coded fields when the reporting source changes. Grouped confusion matrices (Fig 4E,F) illustrate the error structure: under source shift, the code-only model misclassifies bone cement as stent (19%) and mesh as bone cement (13%) at notable rates, while the narrative model maintains high per-class accuracy across all three device classes. Manufacturer reporting profiles underlying the grouped design are shown in S13, and a summary of all quantitative benchmarks is provided in S2.

### Narratives recover interpretable complication phenotypes

Given the greater portability of the narrative representation, the next question is whether it also recovers clinically meaningful complication information in an interpretable way. A binary complication phenotype label was constructed from patient problem codes (Pain, Infection, Hemorrhage, Tissue Injury, Dissection, Erosion, Adhesion, Abscess, Obstruction, or Failure of Implant). Because this label is derived from structured codes rather than independent clinical adjudication, the task is complication phenotype recovery: testing whether narrative text can reconstruct outcome information already present in the coding system and produce an interpretable complication lexicon, not discover outcomes invisible to codes.

Of 1,500 reports, 672 (44.8%) met the complication phenotype definition. Complication prevalence varied substantially by device class (Fig 5A; event type severity context in S14): mesh had the highest rate, driven by Pain and Adhesion codes, consistent with the chronic host-response profile identified in the phenotype analysis, while stent had the lowest rate (22.6%), reflecting its deployment-failure–dominated reporting profile where procedural complications are captured by product problem codes rather than patient problem codes. Across five repeated stratified 80/20 holdouts, logistic regression on TF-IDF features achieved mean F1 = 0.902 (SD = 0.022) and mean AUC = 0.967 (SD = 0.005; Fig 5B), supporting stability across different data partitions (confusion matrix in Fig 5C).

The coefficient profile (Fig 5D) provides the key interpretability result. Positive predictors included patient-outcome and anatomical terms such as “pain,” “dissection,” “infection,” “adhesions,” “knee,” and “hip,” whereas negative predictors included device-identity terms and reporting-template phrases. This separation is consistent with the task definition: the model is recovering a code-derived complication phenotype from narrative language, not predicting an independently adjudicated clinical outcome. The analysis therefore demonstrates that narrative text preserves clinically interpretable outcome signal already present in the structured coding system, which is relevant for regulatory contexts where model transparency is required [31], while also exposing the need for future validation against manually adjudicated narrative labels.

Because the label was derived from patient-problem codes, false positives may include narratives describing clinically plausible complications not captured by the selected code set, whereas false negatives may reflect terse narratives or complication language embedded in reporting-template text. This task is therefore interpreted as phenotype recovery rather than independent outcome discovery. Representative correctly classified reports and edge cases are shown in S6.

### Sparse material naming limits material-specific surveillance from MAUDE narratives alone

Finally, whether explicit material names in MAUDE narratives could support material-aware surveillance was examined. This analysis is exploratory because MAUDE, PubMed, and https://ClinicalTrials.gov differ in scope, time period, reporting incentives, and ascertainment method.

Cross-source material divergence is therefore treated as a prioritization heuristic rather than a source-normalized measure of surveillance coverage or comparative safety. Explicit material naming was rare in MAUDE: only 23 of 1,500 reports (1.5%) named one of the 13 biomaterial classes in the dictionary (Fig 6A). This sparsity suggests that adverse-event narratives typically describe devices in procedural, anatomical, and clinical terms rather than material-composition terms, and that material-specific surveillance will require device-material mapping or domain-specific extraction beyond keyword matching.

**Figure 6.**
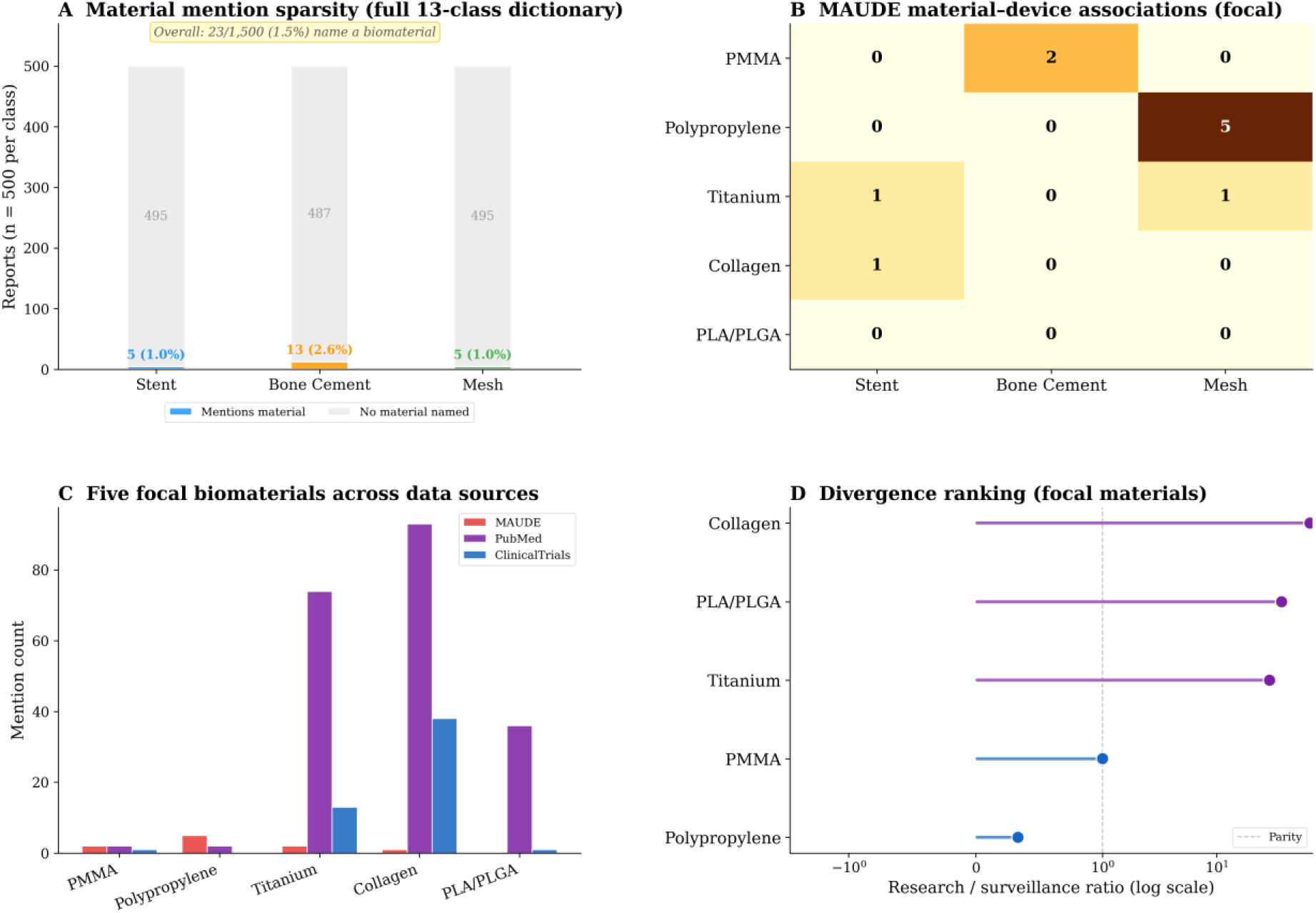
Sparse material naming limits material-specific surveillance from MAUDE narratives alone. (A) Overall material-naming sparsity across the full 13-class dictionary and three device classes (23/1,500 reports, 1.5%). (B) MAUDE material–device heatmap for the five focal materials, showing sparse but expected associations. (C) Grouped comparison of focal-material mentions across MAUDE, PubMed, and https://ClinicalTrials.gov. (D) Divergence ranking ordering focal materials by research-to-surveillance ratio. The cross-source panels are hypothesis-generating and should not be interpreted as comparative safety or risk estimates.

The main cross-source comparison was restricted to five pre-specified biomaterial classes (PMMA, polypropylene, titanium, collagen, and PLA/PLGA), chosen to balance direct cohort relevance and broader translational activity, while retaining the full 13-material scan in the Supporting Information (S17 Fig). Among the five focal materials (Fig 6B), PMMA appeared almost exclusively in bone cement reports and polypropylene in mesh reports, consistent with expected material–device associations. Cross-source comparison (Fig 6C) revealed markedly different visibility profiles: collagen and PLA/PLGA were frequent in the retrieved PubMed and https://ClinicalTrials.gov corpora but were not explicitly named in this MAUDE cohort, whereas PMMA and polypropylene were more visible in surveillance narratives than in the research/trial comparison sets. The research-to-surveillance ratio is therefore interpreted (Fig 6D) as a follow-up prioritization heuristic rather than as evidence of inadequate post-market monitoring or comparative safety. Under this interpretation, titanium and collagen emerge as reasonable candidates for device–material mapping or higher-recall extraction in future surveillance studies. A surveillance-versus-research scatter plot for all 13 material classes is provided in S16. Extended cross-source analyses are in S7–S9.

## Discussion

Across a balanced MAUDE cohort of 1,500 biomaterial-device reports, this study shows that free-text adverse-event narratives provide a more source-robust representation of reported failures than coded fields alone. The principal contribution is not high device-class classification accuracy under ordinary cross-validation, but a representation-level benchmark showing that narrative-derived features retain more information than coded taxonomies when the reporting source changes. Under manufacturer-held-out evaluation, narrative features outperformed code-only structured features (F1 = 0.925 versus 0.827), and delexicalized narratives retained strong grouped performance after masking device, manufacturer, brand, and legal-template terms (F1 = 0.897). These findings support narrative text as a complementary representation layer for post-market device surveillance rather than as a replacement for structured coding.

Representation quality determines what failure patterns surveillance systems can detect under real-world reporting variation, and this portability dimension has been underexplored for device surveillance. Coded signal detection is standardized and scalable, but the observed portability gap suggests that code-based trend analyses may be confounded by changes in manufacturer reporting practices over time. The NMF topic layer provides a second dimension of evidence: after removing legal and administrative boilerplate, several narrative topics capture device-specific clinical content at finer granularity than the coded taxonomy, while others reflect reporting patterns invisible in structured fields. The cross-source material analysis, though exploratory, shows that adverse-event narratives describe devices in procedural and clinical terms rather than materials terms (1.5% explicit material naming), suggesting that material-specific surveillance may require dedicated extraction approaches beyond keyword matching.

Operationally, these results motivate integrating a narrative phenotype layer into surveillance workflows alongside existing coded infrastructure. Structured coding remains essential for scalable signal detection, but narrative representations preserve procedural, anatomical, and complication-related detail that codes compress, and appear less vulnerable to reporting-source conventions. This multi-representation framing is consistent with recent calls for diversified approaches to post-market surveillance [25].

Several limitations bound these conclusions. First, MAUDE is a passive surveillance system; the present study cannot establish incidence, prevalence, causal relationships, or comparative device risk [9,32,33]. Reporting rates vary by manufacturer, event severity, and media attention; duplicate and incomplete reports are common, and these findings describe patterns within reported data, not population-level device safety. Second, the grouped manufacturer-held-out analysis provides a meaningful source-shift stress test, but fold-level class imbalance (narrative fold-level macro F1 ranged from 0.693 to 1.000) means grouped point estimates should be interpreted cautiously; the delexicalized sensitivity analysis partially addresses this concern. Third, the complication phenotype is code-derived rather than externally adjudicated, so the classifier evaluates recovery of clinically meaningful surveillance signal rather than independent outcome discovery; independent clinical adjudication would provide a stronger evaluation. Fourth, the study is limited to three biomaterial device classes, a single annual sampling frame, and a deterministic convenience sample of 500 reports per device class; broader external validation across device types, reporting periods, and sampling strategies remains necessary. Fifth, TF-IDF is a bag-of-words model that discards word order, negation, and context; transformer-based biomedical language models (BioBERT, PubMedBERT) would capture richer semantic representations, though at the cost of interpretability.

Future work should expand the benchmark across additional years and device classes, add temporal validation, and test within-device failure-phenotype tasks that remove broad device vocabulary as a shortcut. Independent clinical adjudication of a stratified narrative subset would strengthen the complication analysis by separating true model error from incomplete or inconsistent coding. More expressive biomedical language models may capture richer contextual information than TF-IDF, but their use should be balanced against transparency requirements in regulatory and patient-safety settings. In summary, free-text MAUDE narratives are not merely richer annotations; they constitute a complementary, source-robust representation layer for characterizing reported biomaterial-device failures under real-world reporting variation.

## Materials and Methods

### Data extraction and preprocessing

Adverse event reports were extracted from the FDA MAUDE database via the openFDA API (https://api.fda.gov/device/event) on March 15, 2026. Three biomaterial device categories were queried using FDA product codes: coronary stents (NIQ, MAF), bone cement (OZO), and surgical mesh (ORC, QFG, QMD). For each category, 500 reports were retrieved from calendar year 2022 by querying with date_received in the range 2022-01-01 to 2022-12-31, sorted by date_received, and taking the first 500 results returned by the API. This fixed-size sequential sampling design was chosen to obtain a reproducible, balanced proof-of-concept cohort across device categories while preserving a common reporting-year window; it is not a random sample of all 2022 reports for each product code and should not be interpreted as representative of all MAUDE reports for these product codes. Extracted fields included: report_number, mdr_text (event_description and manufacturer_narrative, concatenated with event_description first), product_problems, patient_problems, event_type, manufacturer_name (stored as manufacturer_name in the dataset; sourced from the API field device.manufacturer_d_name), brand_name (from device.brand_name), and product_code (from device.product_code). Deduplication was performed on report_number; reports sharing the same report_number (which can occur when follow-up reports update an original filing) were collapsed to the first occurrence to avoid counting the same event multiple times across follow-up filings. MAUDE documentation notes that displayed report content may reflect the most recent follow-up, but the extraction retained the first instance per report_number to avoid double-counting events. No reports were excluded on the basis of report type or exemption status. All reports contained non-empty narrative text; no imputation was required for TF-IDF vectorization.

### Feature engineering

Structured features consisted of one-hot encoded product-problem codes (93 unique values) and patient-problem codes (158 unique values), with narrative word count added only in the structured-plus-word-count sensitivity model, yielding a 252-dimensional feature vector. Because product-problem and patient-problem codes are standardized FDA-coded fields rather than learned text features, the observed code vocabulary was fixed before model fitting; no label information was used in constructing the code matrix. A code-only ablation variant (251 dimensions) excluded word count to isolate the contribution of coded taxonomies alone. Narrative features were generated from the concatenated event_description and manufacturer_narrative text fields using scikit-learn TfidfVectorizer (max_features = 5,000; ngram_range = (1, 2); min_df = 3; stop_words = “english”). For all predictive benchmarks, TF-IDF vocabulary selection and inverse document-frequency weights were fit within each training fold and then applied to the held-out fold, ensuring that test-fold vocabulary and document-frequency information did not influence feature construction. A full-corpus TF-IDF matrix was used only for non-predictive descriptive analyses, including NMF topic display and phrase visualization. Combined features were generated by horizontally concatenating the fold-specific TF-IDF matrix with the structured feature matrix.

### Classification models

Three classifiers were trained using scikit-learn v1.3 (Python 3.10): Logistic Regression (LR; L2 penalty, C = 1.0, max_iter = 1,000, one-vs-rest for multiclass), Random Forest (RF; 200 trees, default hyperparameters), and Gradient Boosting (GB; 200 estimators, max_depth = 5, learning_rate = 0.1). All models used random_state = 42. The primary metric was macro-averaged F1 score (unweighted mean of per-class F1 scores), chosen because class sizes were balanced (500 per category).

### Evaluation protocol

Standard evaluation: 5-fold stratified cross-validation (StratifiedKFold, shuffle = True, random_state = 42). Predictions from all folds were concatenated for metric computation (n = 1,500). Bootstrap 95% confidence intervals: 500 resamples of the cross-validated prediction vector, computing macro F1 on each resample; intervals are the 2.5th and 97.5th percentiles. Manufacturer-grouped evaluation: GroupKFold (5 splits) using manufacturer_name as the grouping variable, ensuring that all reports from any given manufacturer appeared in exactly one fold. Because GroupKFold does not stratify by category, the resulting folds were not balanced by device class (fold composition is reported in S1). This imbalance is an inherent consequence of the manufacturer-grouping design and means that some folds contain disproportionate numbers of one device class. To assess whether category imbalance across folds drives the observed portability gap, a delexicalized narrative representation was additionally tested as a sensitivity analysis (S5). Because GroupKFold preserves manufacturer grouping but does not preserve class balance, both aggregate grouped predictions and fold-level macro F1 are reported; the grouped analysis is interpreted as a reporting-source stress test rather than as a precise estimate of deployment performance. No p-values were computed and no multiple-comparison corrections were applied; this study is primarily descriptive and the confidence intervals serve to quantify estimation uncertainty.

### Complication phenotype classification

A binary complication phenotype label was constructed from patient problem codes. Reports were labelled positive if their patient_problems field contained any of: Pain, Infection, Hemorrhage, Tissue Injury, Vascular Dissection, Erosion, Adhesion, Abscess, Obstruction, or Failure of Implant (672/1,500 = 44.8% positive). This label is derived from structured codes, not independent clinical adjudication. Prediction used TF-IDF features with Logistic Regression (class_weight = “balanced”, C = 1.0, max_iter = 1,000). To assess stability, five repeated stratified 80/20 holdout splits were performed (random_state = 42, 52, 62, 72, 82; training n = 1,200, test n = 300 per repeat). F1 and AUC were computed on each held-out test set and averaged; results are reported as mean ± SD.

### Topic modeling

Non-negative matrix factorization (NMF) was applied to event description text only (excluding manufacturer narratives, which are dominated by template language) after a two-pass cleaning procedure: (1) regex-based removal of legal, litigation, administrative, and MedWatch template sentences (10 patterns), and (2) an expanded stop-word list combining scikit-learn English stop words with 532 additional tokens covering manufacturer names, brand names, regulatory terminology, and reporting boilerplate. The cleaned text was then vectorized with TF-IDF (max_features = 5,000; ngram_range = (1,2); min_df = 3) and factorized with NMF using 8 components (selected after evaluating solutions from k = 5 to k = 15 for interpretability and clinical relevance; S4). This cleaning procedure was applied only to the topic modeling and phenotype display analysis; classification benchmarks used the original unmodified concatenated narrative. Topic stability was assessed by re-running NMF with random_state = 99; matched topic vectors were qualitatively similar across runs (S4). Topic labels were auto-generated from the dominant device class and top informative term per component.

### Material extraction and cross-source analysis

Material mentions were extracted using curated regex patterns for 13 biomaterial classes (PMMA, stainless steel, nitinol, cobalt-chromium, titanium, polyethylene, polypropylene, silicone, collagen, chitosan, PLGA/polylactic acid, silk fibroin, alginate) with case-insensitive matching and word-boundary anchors to prevent partial-word matches. The same 13 patterns were applied to PubMed abstracts (n = 796; retrieved via NCBI E-utilities; query: “biomaterial adverse event”; fields: title and abstract) and https://ClinicalTrials.gov entries (n = 595; retrieved via API v2; query: “biomaterial”; field: brief_title). Five focal biomaterial classes were pre-specified for the main cross-source comparison using three criteria: direct relevance to the sampled device cohort (PMMA for bone cement, polypropylene for mesh), recurrent appearance in the PubMed or https://ClinicalTrials.gov screens (titanium, collagen), and coverage of both permanent and degradable biomaterial families (PLA/PLGA). The complete 13-class dictionary was retained for supplementary sensitivity analysis. Counts were not normalized across sources because the three corpora differ in scope, time period, and ascertainment method; the cross-source comparison is therefore descriptive prioritization rather than comparative quantification. A single report could match multiple material classes; counts were aggregated at the class level (e.g., PLGA and polylactic acid were merged into PLA/PLGA). The 24 material-class matches occurred in 23 unique reports because one report matched two material classes. Regex precision was validated by manual inspection of all 24 MAUDE matches (100% precision, 0 false positives), and recall was assessed by checking 50 sampled non-matching reports per material class for alternate spellings (0 false negatives detected); the word-boundary anchored patterns are therefore conservative (high precision) but may miss non-standard material references. Research-to-surveillance ratio was computed as (PubMed + ClinicalTrials) / (MAUDE + 1) to avoid division by zero.

### Ethics statement

This study exclusively uses publicly available, de-identified adverse event reports from the FDA MAUDE database accessed through the openFDA API. No identifiable patient or provider information was collected. Institutional review board approval was not required because the study does not involve human subjects, identifiable private information, or interventions (45 CFR 46.102).

### Statistical reporting

All metrics are macro-averaged unless otherwise stated. Error bars in figures represent bootstrap 95% confidence intervals (500 resamples). Exact sample sizes are reported for each analysis. No hypothesis tests were performed and no multiple-comparison corrections were applied. All analyses were conducted in Python 3.10 using scikit-learn 1.3, NumPy 1.24, SciPy 1.11, and matplotlib 3.7. Claude (Anthropic) was used as an AI coding assistant during data analysis, figure generation, and manuscript formatting; all scientific content, interpretations, and claims were authored and verified by the human author.

## Acknowledgments

The author received no specific funding for this work. The author thanks the maintainers of the FDA MAUDE/openFDA, PubMed, and https://ClinicalTrials.gov public data resources.

## Author Contributions

Hongyi Chen: Conceptualization, Methodology, Software, Data curation, Formal analysis, Visualization, Writing – original draft, Writing – review & editing.

## Data Availability Statement

No new primary data were generated. The following existing public data sources were used: FDA MAUDE adverse-event reports from the openFDA API (https://open.fda.gov/apis/device/event/), PubMed abstracts retrieved via NCBI E-utilities (https://eutils.ncbi.nlm.nih.gov/), and https://ClinicalTrials.gov entries retrieved via API v2 (https://clinicaltrials.gov/data-api/). Processed datasets, analysis scripts, and generated result files will be deposited in a public repository (GitHub/Zenodo) with a DOI prior to publication.

## Supporting Information

S1–S17 Figs and S1–S4 Tables (numbered in order of first citation) are provided as Supporting Information.

